# Evaluation of genetic associations with clinical phenotypes of kidney stone disease

**DOI:** 10.1101/2024.01.18.24301501

**Authors:** Ryan S Hsi, Siwei Zhang, Jefferson L Triozzi, Adriana M Hung, Yaomin Xu, Cosmin A Bejan

## Abstract

**Introduction and Objective:** We sought to replicate and discover genetic associations of kidney stone disease within a large-scale electronic health record (EHR) system.

**Methods:** We performed genome-wide association studies (GWASs) for nephrolithiasis from genotyped samples of 5,571 cases and 83,692 controls. Among the significant risk variants, we performed association analyses of stone composition and first-time 24-hour urine parameters. To assess disease severity, we investigated the associations of risk variants with age at first stone diagnosis, age at first procedure, and time from first to second procedure.

**Results:** The main GWAS analysis identified 10 significant loci, each located on chromosome 16 within coding regions of the *UMOD* gene, which codes for uromodulin, a urine protein with inhibitory activity for calcium crystallization. The strongest signal was from SNP 16:20359633-C-T (odds ratio [OR] 1.17, 95% CI 1.11-1.23), with the remaining significant SNPs having similar effect sizes. In subgroup GWASs by stone composition, 19 significant loci were identified, of which two loci were located in coding regions (brushite; *NXPH1*, rs79970906 and rs4725104). The *UMOD* SNP 16:20359633-C-T was associated with differences in 24-hour excretion of urinary calcium, uric acid, phosphorus, sulfate; and the minor allele was positively associated with calcium oxalate dihydrate stone composition (p<0.05). No associations were found between *UMOD* variants and disease severity.

**Conclusions:** We replicated germline variants associated with kidney stone disease risk at *UMOD* and reported novel variants associated with stone composition. Genetic variants of *UMOD* are associated with differences in 24-hour urine parameters and stone composition, but not disease severity.

## Introduction

Multiple lines of evidence support a strong genetic contribution to kidney stone risk, including familial studies, twin studies, and studies on single-gene mutations.^1–6^ In addition, prior large-scale genome-wide association studies (GWASs) from the UK, Japanese and Icelandic populations have implicated genetic variants linked to calcium and phosphate regulation, metabolic traits, and inflammation/oxidative stress.^7–12^ Overall, previous studies have reported that kidney stone risk is approximately 50% attributable to genetic heritability.^2,3^

Despite such a large contribution of genetics to stone risk, clinicians uncommonly utilize genetic information for clinical care for several reasons. First, monogenic causes for kidney stones are uncommon.^13^ At present, genetic screening is not routinely performed except for primary hyperoxaluria or cystinuria, and often, these conditions can be inferred indirectly from stone analysis and urinary levels of cystine and oxalate. Furthermore, while many genetic studies have elucidated mechanisms for disease risk in population-based epidemiology studies, few have examined those associations with clinical phenotypes such as disease severity in patient clinical context– only one GWAS has identified rare variants associated with recurrent kidney stones.^7^ As such, there is an unmet need to evaluate genetic factors in real-world patient populations, interpret genetic factors linked to patient clinical outcomes, and assess how well these genetic factors may fit into existing treatment algorithms.

Within this context, we investigated the translational potential of using genome-wide association findings for risk prediction and disease subclassification. First, we performed a GWAS for kidney stone disease within an electronic health record framework (EHR). In a subgroup analysis, we performed separate GWASs among subgroups of individuals with kidney stones classified by stone composition. Then, for SNPs meeting the criteria for genome-wide significance, we compared differences in 24-hour urine values and stone composition by allele status, and evaluated associations with disease severity.

## Methods

### Data source and study population

We investigated a de-identified version of the entire electronic health record from our single institution (the Synthetic Derivative, SD), which is updated bimonthly and contains longitudinal clinical records of 3.2 million records since 1982. The SD is linked to a biobank (BioVU)^14^ which has accrued DNA samples since 2007 from unused blood drawn for routine clinical practice scheduled to be discarded. Our study population included individuals within the SD with genotyping data from BioVU (n=90,991). Local institutional review board approval was obtained for this study with waiver of consent (IRB# 190480).

### Kidney stone cases and controls

We identified kidney stone cases within the SD using Current Procedural Terminology (CPT) codes and International Classification of Diseases, 9th/10th Revision, Clinical Modification (ICD-9/10-CM) diagnosis codes within the SD up to May 2021. We required a single ICD or CPT code to be classified as a case (see Appendix 1 for relevant inclusion codes for kidney stone-related diagnoses). Individuals without any of these codes were considered non-cases. To ensure these codes reflected kidney stone cases, a manual review of records containing kidney stone-related ICD codes (n=200) and CPT codes (n=200) showed PPVs of 93% and 97%, respectively. Then, among individuals identified with kidney stone disease, we identified comorbid conditions prior to the index kidney stone diagnosis based on ICD codes listed in Appendix 2.

Controls were identified from non-cases who did not have additional diagnosis codes for exclusion (see Appendix 1). These exclusion codes included diagnoses of hydronephrosis and lower urinary tract stones.

**Appendix 1.**
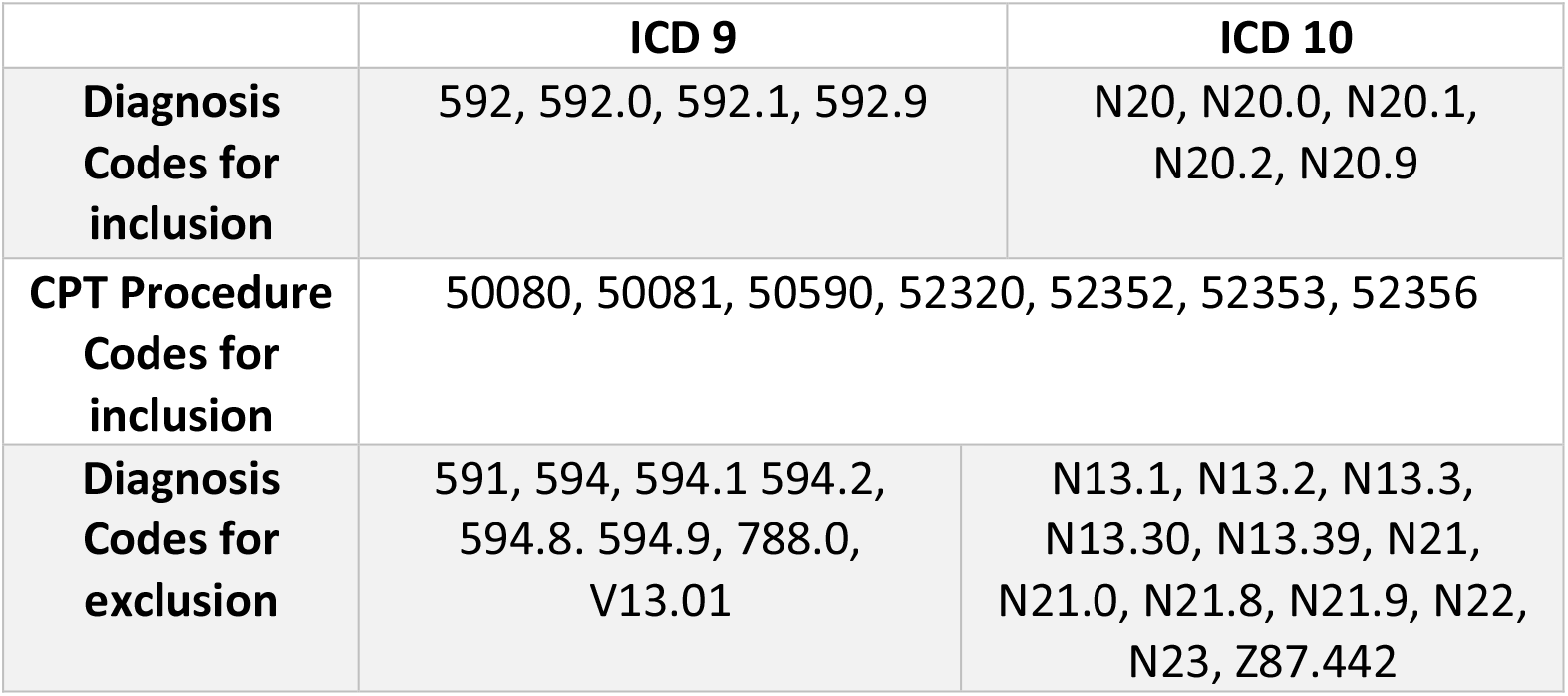
ICD and CPT codes for kidney stone disease.

|  | **ICD 9** | **ICD 10** |
| --- | --- | --- |
| **Diagnosis Codes for inclusion** | 592, 592.0, 592.1, 592.9 | N20, N20.0, N20.1, N20.2, N20.9 |
| **CPT Procedure Codes for inclusion** | 50080, 50081, 50590, 52320, 52352, 52353, 52356 |  |
| **Diagnosis Codes for exclusion** | 591, 594, 594.1 594.2, 594.8. 594.9, 788.0, V13.01 | N13.1, N13.2, N13.3, N13.30, N13.39, N21, N21.0, N21.8, N21.9, N22, N23, Z87.442 |

### Genotyping and imputation

Genotyping of BioVU samples was performed with the Illumina Expanded Multi-Ethnic Genotyping Array (MEGA^EX^), which targets almost 2 million exome and rare single nucleotide polymorphisms (SNPs). The exome chip processing protocol and quality control procedures have been previously described.^15^ We performed standard quality control procedures to exclude low-quality variants and individuals including SNPs with missingness >2%, individuals with missingness >5%, SNPs with minor allele frequency (MAF) < 1% and p-value of Hardy-Weinberg equilibrium test of < 1e-6^16^. Then, the data were imputed to the 1000 Genomes Project Phase 3 reference panel^17^ for haplotype estimation and imputation. We converted dosage data to hard genotype calls and excluded variants with uncertainty > 0.1 or INFO < 0.95, resulting in 2,225,361 variants after post-imputation quality control.

### GWAS

In the GWAS, the association of SNPs with kidney stone disease was performed using logistic regression with an additive genetic model and adjusting for age at diagnosis, race (White, Black, Other), sex, and 10 principal components to correct for population stratification (see Figure 1).^18,19^ Additive genetic models assessed the linear increase in risk for each copy of the minor allele. A p-value <5x10^−8^ was used for genome-wide significance. We additionally performed two sensitivity analyses with more stringent exclusion criteria defining cases and controls using additional conditions (1) at least 2 years of EHR follow-up data between the timestamp of the first record and last record, and 2) non-missing demographic information (sex, race, ethnicity). Local linkage disequilibrium (LD) and recombination patterns were accessed using LocusZoom^20^. To isolate the independent signals, we performed a conditional analysis in PLINK by adding the lead SNP to the covariates and rerunning the association test. We utilized ANNOVAR for SNP annotation to identify and elucidate the functional implications of genetic variations in the surrounding genome regions.^21^

**FIGURE 1.**
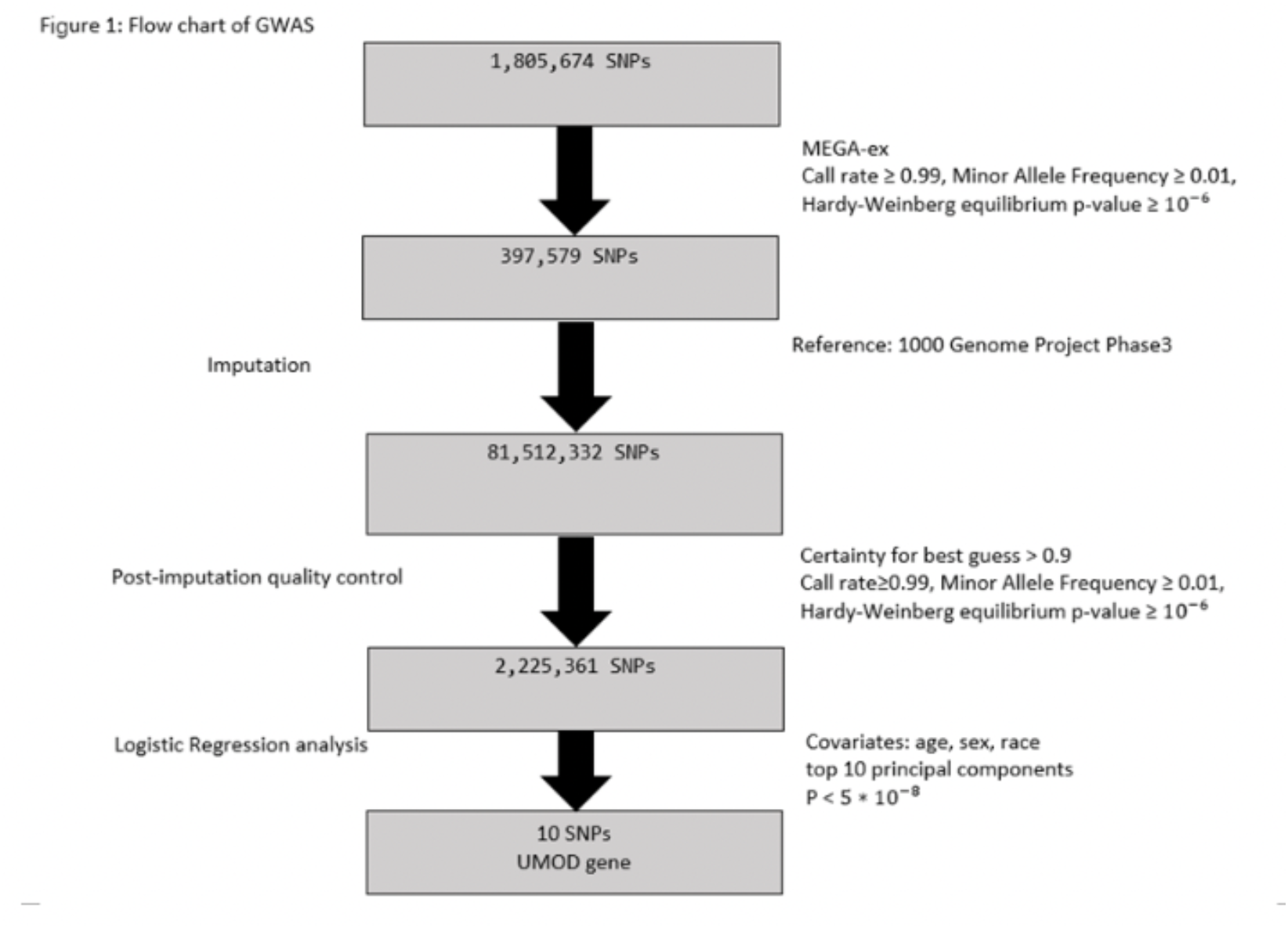
Flow chart of GWAS.

### Subgroup GWASs by stone composition

For genotyped individuals with kidney stone history, the first stone composition (Beck Analytical Services, Greenwood, Indiana) from the kidney or ureter, was identified. Based on classifications described previously,^22^ we identified those with majority calcium oxalate (monohydrate and/or dihydrate), majority calcium oxalate monohydrate, majority calcium oxalate dihydrate, majority hydroxyapatite, any uric acid, any brushite, any carbonate apatite, and any struvite. Separate GWASs were performed for each of the stone composition categories, from which individuals were considered cases. Controls were individuals without kidney stone-related diagnoses or procedure codes, as in the main analysis. The GWAS analyses were then performed as described for the main GWAS study.

### Comparison of stone composition and 24-hour urine

For the significant risk alleles identified in the main GWAS, individuals were assigned to three groups based on allele status. Stone composition (as described previously) and first-time 24-hour urine samples (Litholink, Itasca, IL) were identified among those with kidney stone history. We included only 24-hour urine samples with creatinine/kilogram values within reference ranges (male 11.9-24.4 mg/kg, female 8.7-20.3 mg/kg) to ensure adequate sample collection.

Descriptive statistics and comparisons were performed among the allele groups. For the stone composition analysis, individuals were categorized into mutually exclusive groups of majority calcium oxalate monohydrate, majority calcium oxalate dihydrate, majority hydroxyapatite, and majority uric acid. Logistic regression models were performed for each stone composition category under three different genetic models: dominant, recessive model, and additive models. For each of the 24-hour urine parameters, univariate analysis was performed with ANOVA, and p-values <0.05 were considered significant.

### Disease severity analysis

We assessed the effect of SNP allele status on disease severity for 3 outcomes: 1) age at first kidney stone diagnosis; 2) age at first surgical procedure; and 3) time to second surgery from first surgical procedure. The first two outcomes were evaluated with ANOVA with p-values <0.05 considered significant. To account for the cohort effect associated with the variation among different cohorts at the same age,^23^ the SNPs with significant results from the Kruskal-Wallis test were evaluated in a linear mixed effects model with sex, race, and ethnicity as fixed effects, and birth cohort (every 10 years) as a random effect. Finally, the third outcome was assessed using a time to event analysis with cox proportions hazards regression model adjusting risk factors of sex, race, and ethnicity; the log-rank test was used to compare Kaplan-Meier survival curves and the log-rank test p-values <0.05 were considered significant. A Kaplan-Meier plot was generated to illustrate differences in surgical recurrence probability.

To account for the potential lack of power for the disease severity analyses, we performed equivalence tests for each of the three outcomes. Individuals were split into two groups with one group having at least one minor risk allele and another group without any minor risk allele. The effect size was converted by the difference in the outcome between the two groups given the sample sizes and a pooled standard deviation.

### Software

For the GWAS quality control and association analysis, PLINK 1.9^24^ was used. For the GWAS imputation procedure, IMPUTE2^25^ and SHAPEIT^26^ were used. All the other statistical analysis was implemented in R version 4.1.0.

## Results

Among 89,533 genotyped individuals, the analysis included 5,571 (6.2%) with kidney stone disease and 83,692 (93.8%) controls. Descriptive statistics for the kidney stone cohort are shown in Supplementary Table 1. The mean age at diagnosis was 52.0 years, 51.3% males, and comprised of 86.4% White and 10.5% Black individuals (see Table 1). The most common comorbid conditions were hypertension (52.1%), obesity (24.5%), diabetes type 2 (23.4%), and cardiovascular disease (23.0%).

**Supplementary Table 1:** Demographic and clinical characteristics of genotyped individuals with kidney stone disease.

|  | Kidney stone diagnosis (n=5571) |
| --- | --- |
| Age at diagnosis (years, mean, SD) | 52.0, 18.0 |
| Male gender | 2858 (51.3%) |
| **Race** |  |
| White race | 4812, (86.4%) |
| Black | 585 (10.5%) |
| Other/unknown | 174 (3.1%) |
| **Ethnicity** |  |
| Not Hispanic/Latinx | 5422 97.3% |
| Cardiovascular disease | 1281 23.0% |
| Diabetes Type 2 | 1305 23.4% |
| Gout | 325 5.8% |
| Hypertension | 2906 52.1% |
| Inflammatory bowel disease | 575 10.3% |
| Obesity | 1112 20.0% |

**Table 1.**
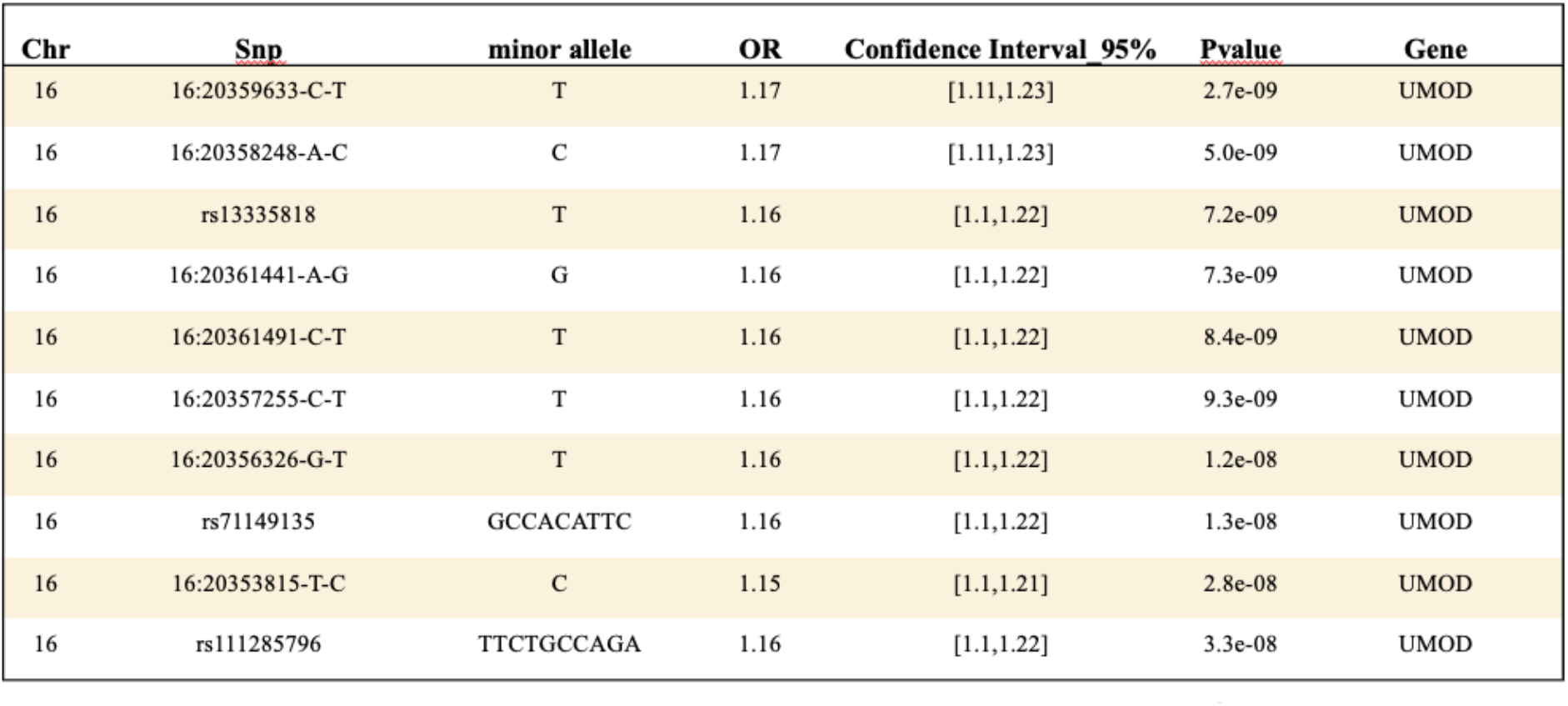
Genetic variants of kidney stone disease meeting genome-wide significance.

| Chr | Snp | minor allele | OR | Confidence Interval 95% | Pvalue | Gene |
| --- | --- | --- | --- | --- | --- | --- |
| 16 | 16:20359633-C-T | T | 1.17 | [1.11,1.23] | 2.7e-09 | UMOD |
| 16 | 16:20358248-A-C | C | 1.17 | [1.11,1.23] | 5.0e-09 | UMOD |
| 16 | rs13335818 | T | 1.16 | [1.1,1.22] | 7.2e-09 | UMOD |
| 16 | 16:20361441-A-G | G | 1.16 | [1.1,1.22] | 7.3e-09 | UMOD |
| 16 | 16:20361491-C-T | T | 1.16 | [1.1,1.22] | 8.4e-09 | UMOD |
| 16 | 16:20357255-C-T | T | 1.16 | [1.1,1.22] | 9.3e-09 | UMOD |
| 16 | 16:20356326-G-T | T | 1.16 | [1.1,1.22] | 1.2e-08 | UMOD |
| 16 | rs71149135 | GCCACATTC | 1.16 | [1.1,1.22] | 1.3e-08 | UMOD |
| 16 | 16:20353815-T-C | C | 1.15 | [1.1,1.21] | 2.8e-08 | UMOD |
| 16 | rs111285796 | TTCTGCCAGA | 1.16 | [1.1,1.22] | 3.3e-08 | UMOD |

The GWAS results show 10 distinct association signals meeting the genome-wide significance, all located in the *UMOD* gene region between 20,344,373 bp-20,364,037 bp (GRCh37) on chromosome 16 (Table 1). The Manhattan plot from the GWAS is shown in Figure 2. SNP 16:20359633-C-T had the strongest signal with minor allele positively associated with higher risk compared to those without the same allele (OR 1.17, 95% CI 1.11-1.23). The conditional analysis indicated that the SNP 16:20359633-C-T represents an independent signal, with the associations of all other SNPs vanishing upon adjustment for this specific SNP. There were two additional variants, 16:20353049-G-A and 16:20359267-G-T within the region of *UMOD* that approached but did not reach the significance threshold. As shown in the LocusZoom and QQ plots based on the UMOD gene region (16p12.3) depicted in Figure 3a and Figure 3b, the associated region and the significant SNPs are in strong LD with the index SNP (r^2^≥0.8). For the two sensitivity analyses restricting case criteria to require 2 years of EHR follow-up data and with complete demographic information, the findings were unchanged.

**Figure 2.**
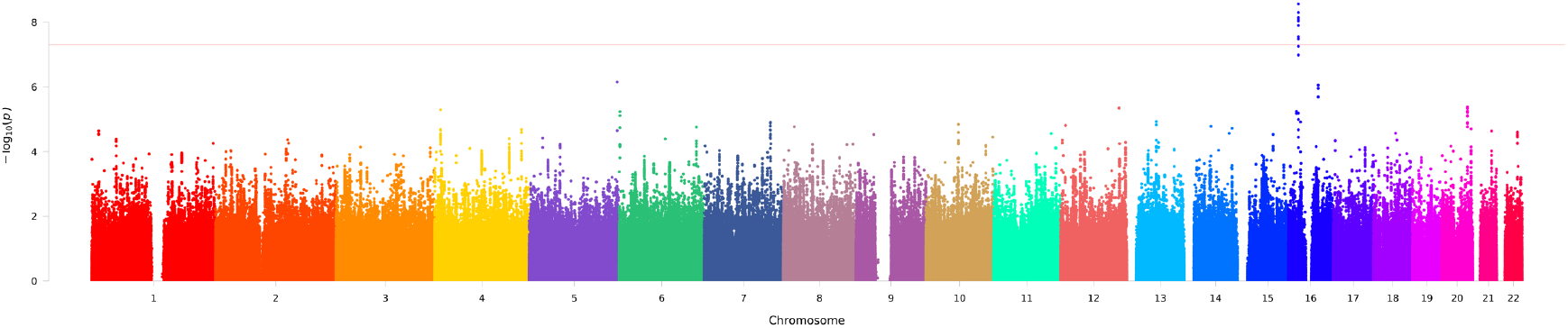
Manhattan plot for the GWAS for kidney stone disease

**Figure 3a.**
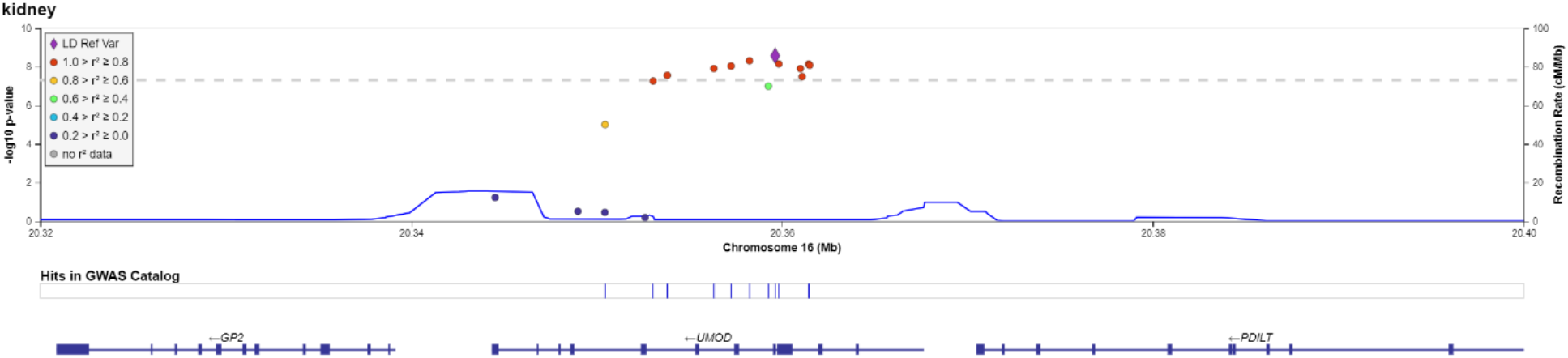
LocusZoom depicting GWAS association data in the context of chromosome 16 in the *UMOD* region.

**Figure 3b.**
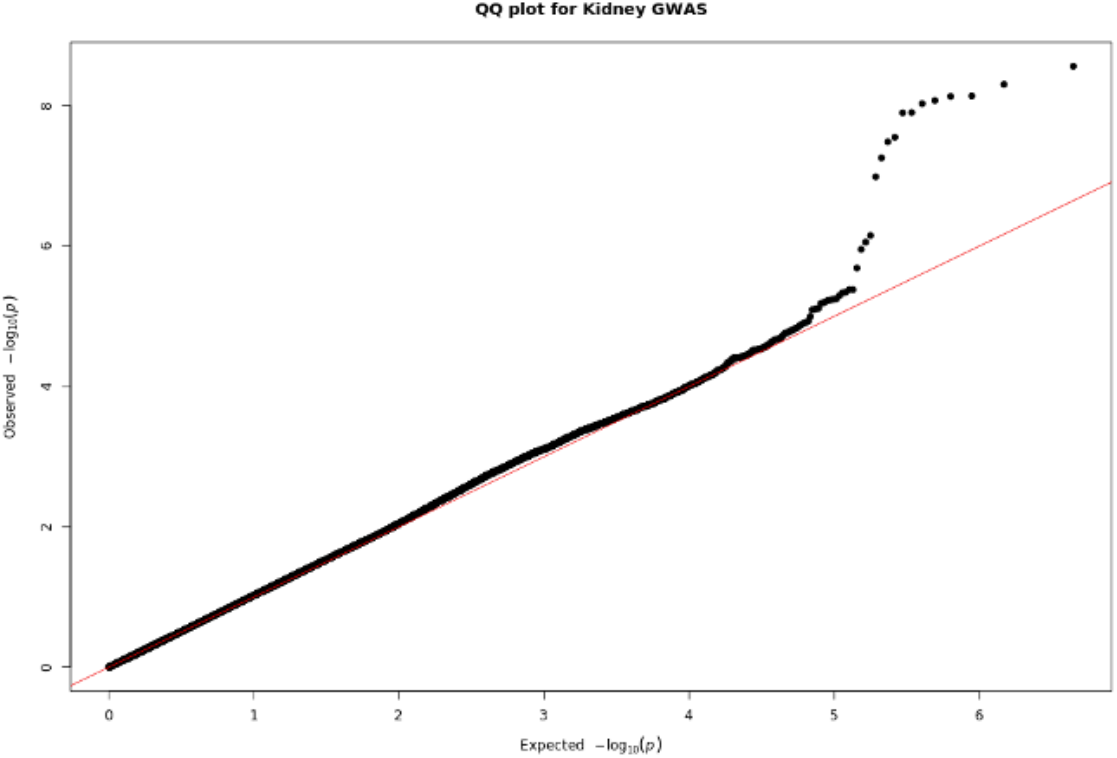
Quantile-Quantile (QQ) plot showing a subset of variants showing association with kidney stone disease.

In the GWASs by kidney stone composition subtypes, we identified 19 loci meeting the threshold for genome-wide significance (see Table 2, Manhattan plots are shown in Supplementary Figure 1). Of these loci, 16 SNPs in 7 genomic regions were located in intergenic regions associating with majority calcium oxalate dihydrate, hydroxyapatite, any uric acid, brushite, carbonate apatite, and struvite. We identified a significant association for brushite stone composition with rs79970906 and rs4725104 in the gene region of *NXPH1* from chromosome 7. Additionally, for struvite stone composition, our study identified a significant association with SNP JHU_8.3486048 on chromosome 8 in a genomic region related to epigenetic function (non-coding RNA LINC01288). Notably, this signal was near the significant locus rs186944649 for carbonate apatite on chromosome 8 in an intergenic position between DUSP26 and LINC01288.

**Table 2.**
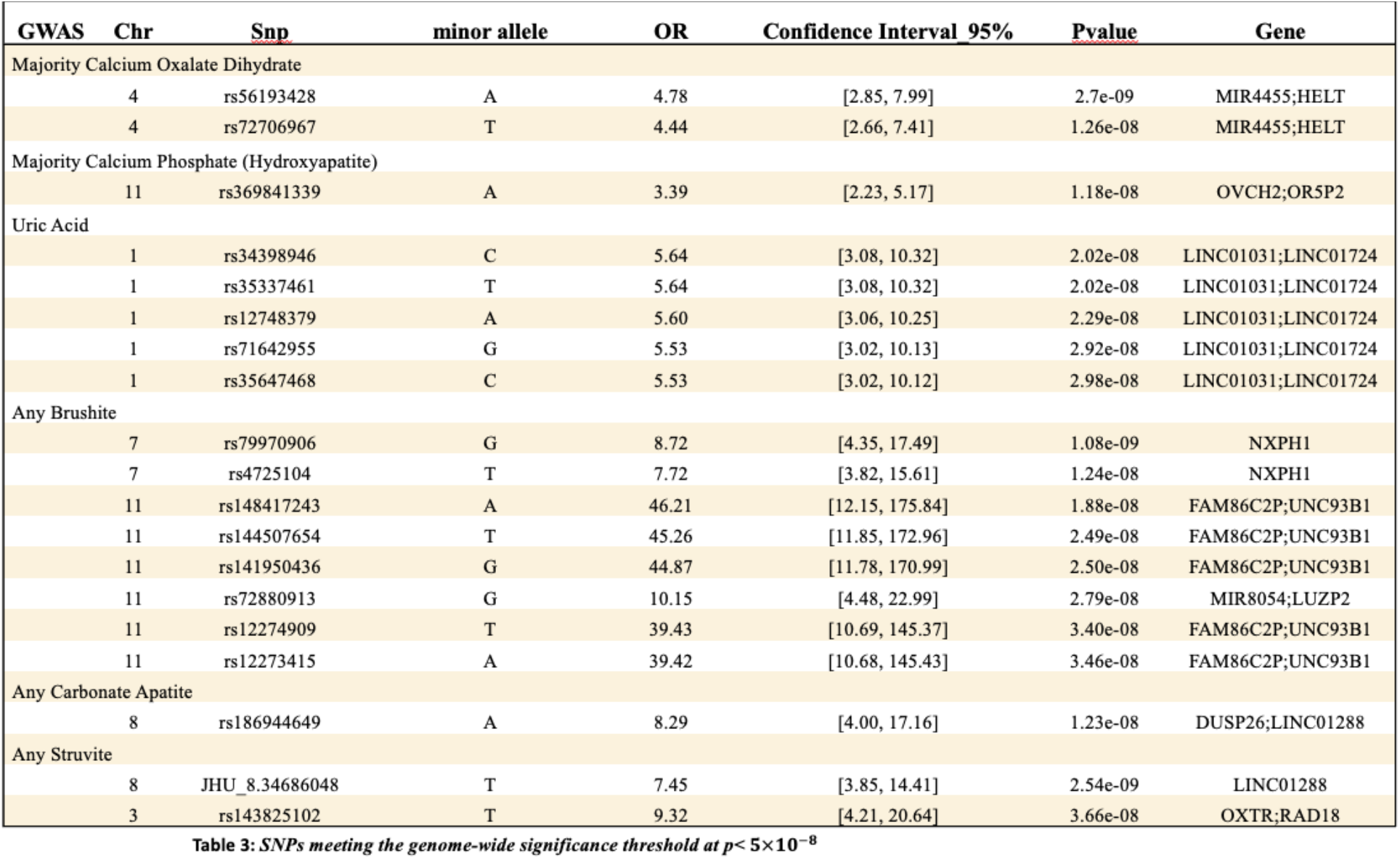
Genetic variants meeting genome-wide significance by kidney stone composition.

Stone composition and 24-hour urine parameters were compared by allele status for SNP 16:20359633-C-T, which had the strongest signal in the main GWAS analysis (see Table 3). Among 586 individuals receiving first-time 24-hour urine testing, values significantly differed for 24-hour urine calcium, uric acid, phosphorus, sulfate (each p<0.05), but not urine supersaturations for calcium oxalate, calcium phosphate, or uric acid (each p>0.05). Among 1,743 individuals with stone composition data, there was a positive association with SNP allele status and majority calcium oxalate dihydrate stone type under either an additive model or dominant model (p<0.05).

**Table 3.** SNP 16:20359633-C-T allele status with 24-hour urine and stone composition.

| 24hr urine parameter,<br>Mean (SD) | No allele<br>(N=376) | 1 allele<br>(N=182) | 2 alleles<br>(N=28) | P-value |
| --- | --- | --- | --- | --- |
| Volume (L) | 1.85 (0.88) | 1.88 (0.88) | 1.77 (0.77) | 0.784 |
| Calcium (mg) | 183 (122) | 209 (135) | 197 (157) | **0.024** |
| Oxalate (mg) | 37.8 (18.2) | 38.4 (18.4) | 34.1 (14.8) | 0.709 |
| Citrate (mg) | 522 (364) | 554 (389) | 435 (269) | 0.345 |
| pH | 6.15 (0.56) | 6.08 (0.54) | 6.24 (0.49) | 0.157 |
| Uric Acid (g) | 0.573 (0.244) | 0.618 (0.253) | 0.559 (0.252) | **0.044** |
| Sodium (mEq) | 166 (81) | 174 (82) | 155 (75) | 0.258 |
| Potassium (mEq) | 56 (26) | 57 (28) | 55 (29) | 0.584 |
| Magnesium (mg) | 94 (45) | 98 (51) | 89 (47) | 0.351 |
| Phosphorus (g) | 0.876 (0.391) | 0.953 (0.401) | 0.866 (0.350) | **0.031** |
| Ammonium (mEq) | 33 (18) | 34 (17) | 35 (27) | 0.460 |
| Chloride (mEq) | 160 (77) | 165 (78) | 147 (70) | 0.487 |
| Sulfate (mEq) | 34 (18) | 38 (20) | 34 (19) | **0.023** |
| Creatinine/kg<br>(mg/kg) | 13.6 (2.8) | 14.1 (2.8) | 12.7 (2.8) | 0.092 |
| SSCaOx | 6.6 (3.9) | 7.1 (4.01) | 6.3 (3.3) | 0.184 |
| SSCaP | 1.3 (1.2) | 1.3 (1.1) | 1.6 (1.3) | 0.607 |
| SSUA | 0.82 (0.87) | 0.96 (0.92) | 0.69 (0.78) | 0.085 |
| Stone composition,<br>% | No allele<br>(N=1109) | 1 allele<br>(N=551) | 2 alleles<br>(N=83) | P-value* |
| Majority CaOxM | 31.9 | 32.3 | 34.9 | 0.262 |
| Majority CaOxD | 5.8 | 8.0 | 8.4 | **0.034** |
| Majority CaPhos | 13.0 | 12.3 | 10.8 | 0.836 |
| Majority Uric Acid | 3.5 | 2.7 | 2.4 | 0.422 |

For the disease severity analysis, we considered SNP 16:20359633-C-T as the representative SNP. The median age at first kidney stone diagnosis was higher among the homozygous major allele (54.2 years) compared to the heterozygous group (53.1 years, p=0.005) and homozygous minor allele group (51.5 years, p=0.03) (see Supplementary Figure 1). However, our analysis for the age at first kidney stone analysis showed no significant association with SNP 16:20359633-C-T (p=0.35) (Supplementary Figure 2). Equivalence testing showed sufficient power to detect a minimum difference of 1 month.

Among those receiving any kidney stone surgery (n=1,106), no differences were observed comparing the age at first surgery, or time to 2^nd^ surgery by 5 years of follow-up (Supplementary Figure 3-5). Equivalence testing showed sufficient power to defect a minimum of 1 month for the age at first surgical procedure outcome, and a minimum of 2 months for the first to second surgery outcome.

**Supplementary Figure 1.**
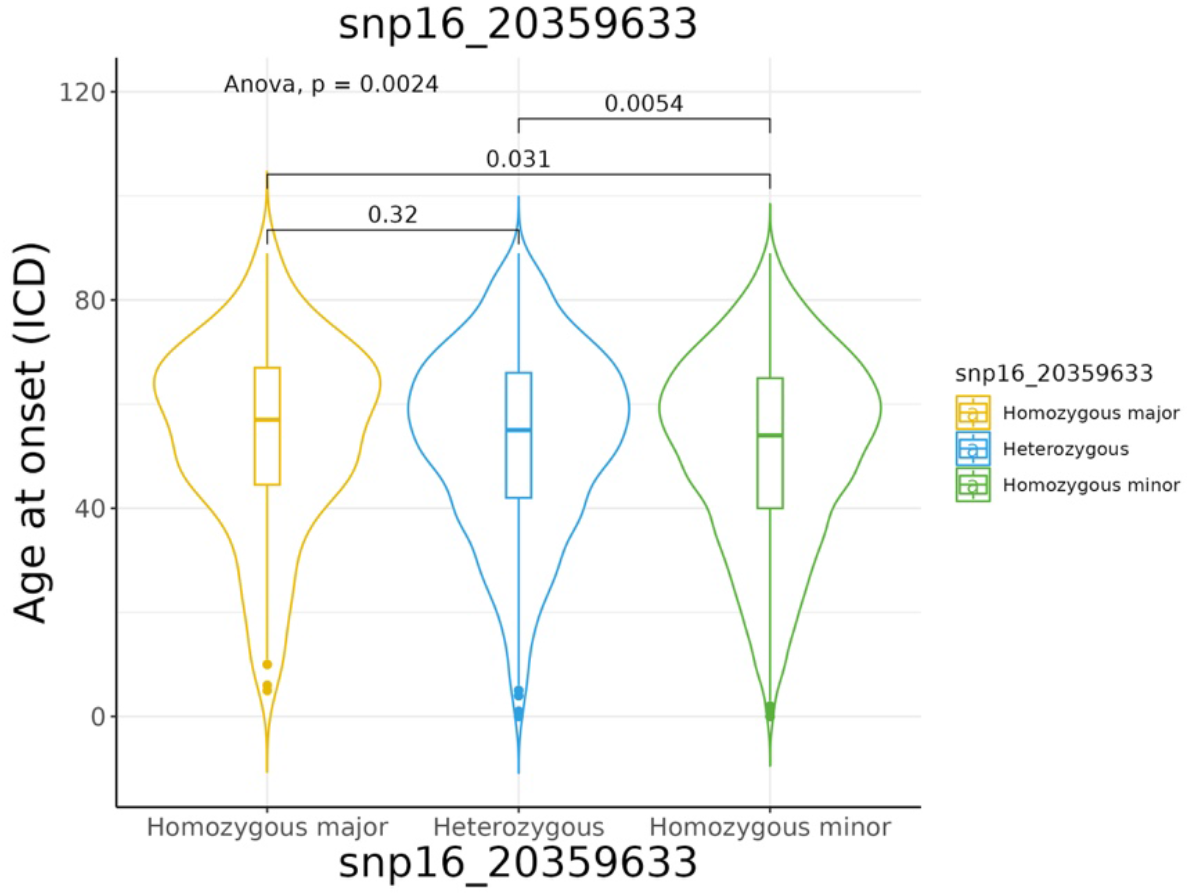
Age at first diagnosis based on SNP 16:20359633-C-T allele status.

**Supplementary Table 2.** Linear mixed effects model for age at first kidney stone diagnosis for SNP 16:20359633-C-T with sex, race, and ethnicity as fixed effects, and birth cohort (every 10 years) as a random effect.

|  | Estimate | Std. Error | df | t value | Pr(>\|t\|) |
| --- | --- | --- | --- | --- | --- |
| (Intercept) | 37.96 | 9.22 | 11.57 | 4.12 | 0.00 |
| snp16_20359633 | -0.29 | 0.32 | 973.02 | -0.90 | 0.37 |
| SexM | -0.42 | 0.38 | 973.11 | -1.09 | 0.28 |
| RaceB | 1.99 | 1.87 | 972.98 | 1.07 | 0.29 |
| RaceW | 2.05 | 1.76 | 972.98 | 1.17 | 0.24 |
| EthnicityHL | 1.00 | 3.25 | 972.98 | 0.31 | 0.76 |
| EthnicityNH | 1.46 | 3.07 | 972.98 | 0.48 | 0.63 |

**Supplementary Figure 3.**
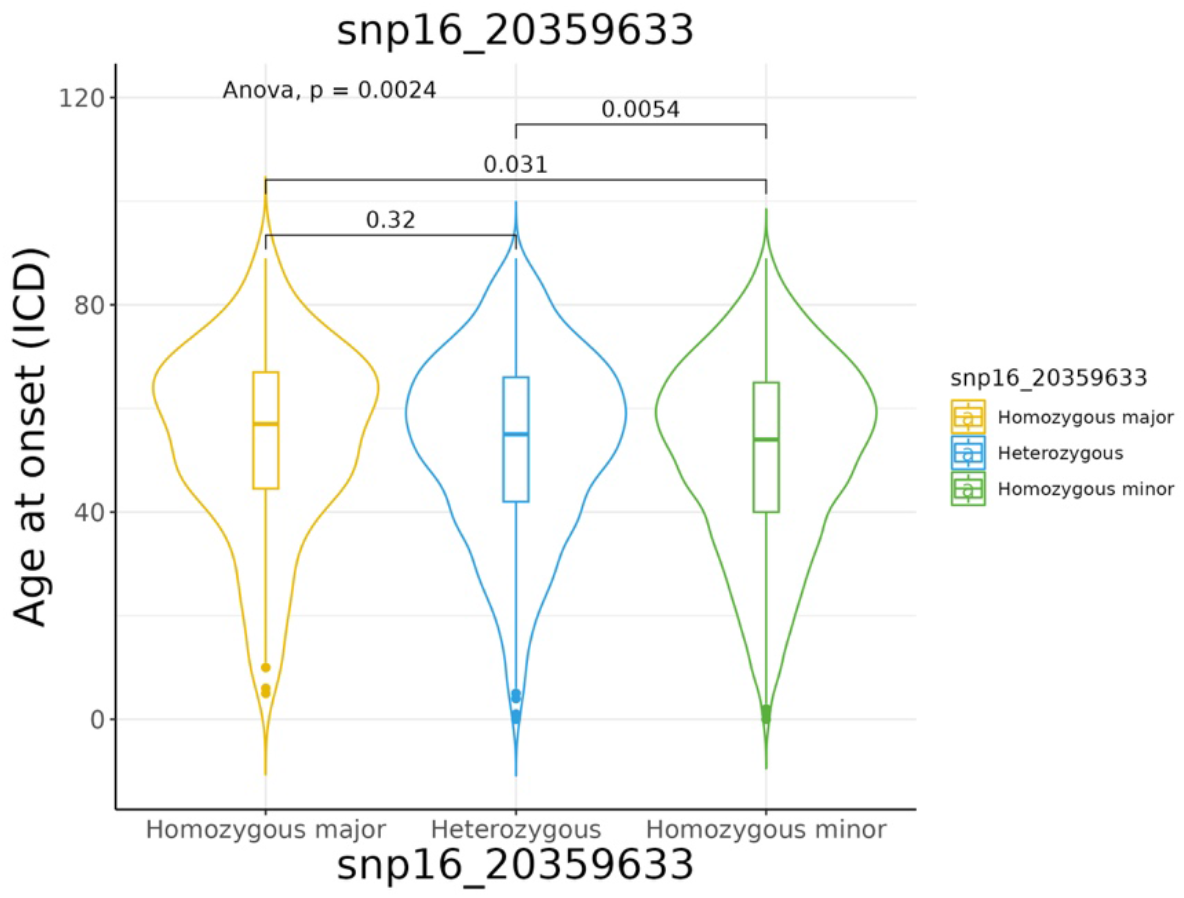
Age at first stone-related surgery based on SNP 16:20359633-C-T allele status.

**Supplementary Figure 4.**
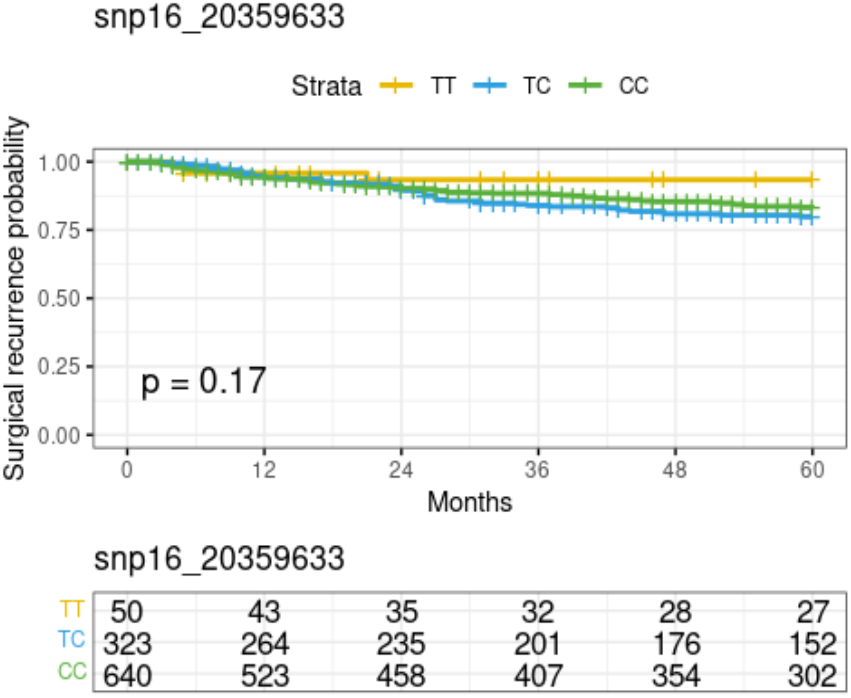
Kaplan-Meier curve showing freedom from second stone-related surgery after first surgery by SNP 16:20359633-C-T allele status. CC and TT are homozygous major and minor, respectively.

**Supplementary Figure 5.**
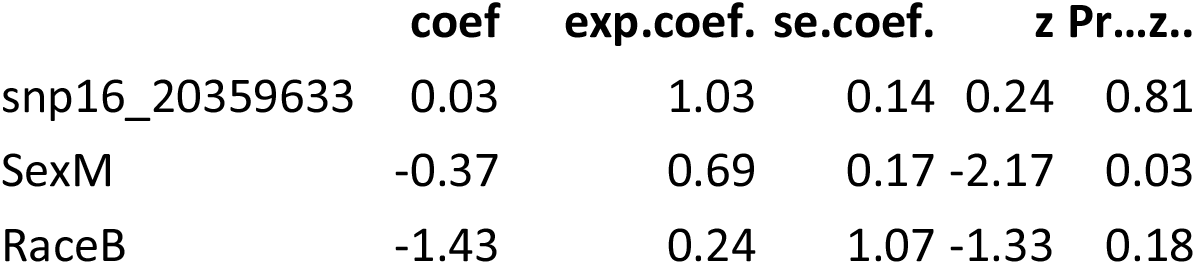
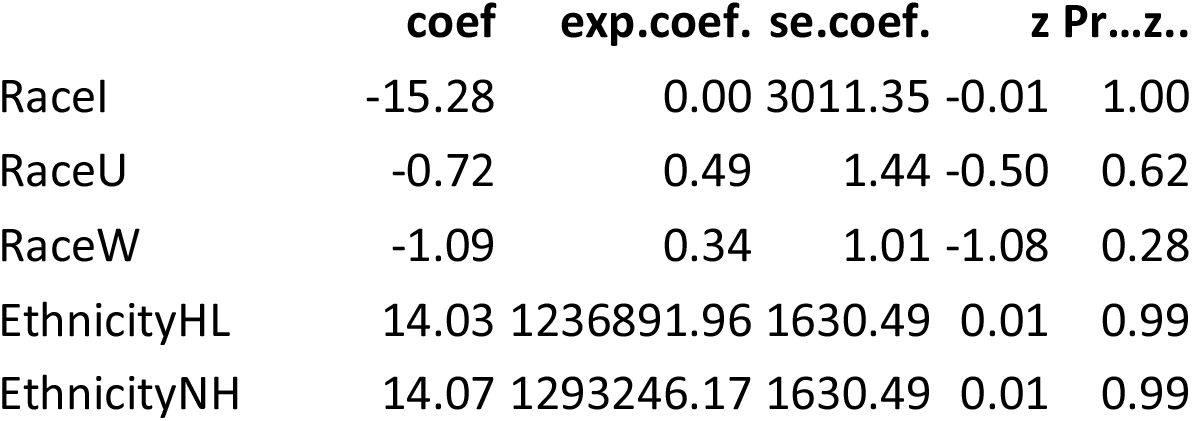
Cox proportions hazards regression model for up to 60 months for the hazard of 2^nd^ stone-related surgery after the 1^st^ stone-related surgery including the SNP 16:20359633-C-T allele status.

## Discussion

We identified several important key findings. First, our results provide an independent replication of multiple risk loci within *UMOD* (16p12.3) in high linkage disequilibrium associating with kidney stone disease risk. *UMOD* encodes uromodulin – also known as Tamm-Horsfall protein, which is the most abundant protein in normal urine and an inhibitor of calcium crystallization^27,28^. In subgroup analysis, we implicate several genes meeting GWAS level of significance associating with stone composition. We additionally demonstrate differences in stone composition and 24-hour urine parameters consistent with uromodulin function as an inhibitor for calcium-based stone types. Furthermore, no relationships with kidney stone severity were identified with *UMOD* risk alleles despite adequate statistical power for the three severity measures evaluated.

Consistent with other GWASs in British, Icelandic and Japanese populations^10–12^, our finding of a positive association between the *UMOD* minor allele variant and kidney stone risk in this U.S. based cohort has a similar mild effect size. In addition to their role in urinary stone risk, *UMOD* variants have been implicated in urinary acidification, renal function, and the development of CKD.^29^ Additionally, common variants in the UMOD promoter region influence urinary uromodulin levels,^30^ suggesting that these SNPs may be potentially causal. The finding of association with calcium oxalate dihydrate stones, which have been linked to urinary calcium excretion,^31^ provides additional supportive evidence. However, our finding of the lack of association between *UMOD* risk alleles and kidney stone severity suggests that while uromodulin influences kidney stone risk, it may not influence subsequent meaningful clinical outcomes. Interestingly, a prior study recurrent kidney stone formers in Iceland, rare missense variants in the sodium-phosphate co-transporter *SLC34A1* and the calcium channel *TRPV5* associated with recurrence risk.^7^ In the current analysis, we tested different measures of disease severity, and work is ongoing to identify and validate stone recurrence episodes within the EHR.

From the 19 novel genome-wide significant loci for kidney stone composition subtypes, we observed that 16 of the loci were located in intergenic regions and one loci (JHU_8.3486048) was located in a non-coding RNA region. While these loci suggest a potential role of regulatory function of the DNA sequence and epigenetics, implicating disease pathways from these SNPs is a challenge. The remaining two loci rs79970906 and rs4725104 show a very strong signal between brushite stone disease and the coding region of *NXPH1. NXPH1* codes for Neurexophilin 1, a secreted protein implicated in irritable bowel syndrome^32^ and several neurologic conditions including Alzheimer’s Disease.^33^ Notably, NXPH1 mediates hematopoiesis, immune responses, and osteoblast activity in the bone marrow,^34^ which could be a plausible pathway to explain the common finding of hypercalciuria in this population.^35^

There are several limitations to this study. EHR-based datasets are susceptible to omitted variable bias and misclassification bias. To validate our method for identifying cases, we performed a manual review of a subset of ICD and CPT codes and demonstrated high PPV for kidney stone diagnoses. As this was an EHR-wide study from a single health care network, we do not capture care sought outside of our health system and would not have identified, for example, surgical procedures at another facility. In addition, these data from the Southeastern U.S. may not be generalizable to other populations.

Notwithstanding these limitations, our study has important clinical implications. Our findings suggest a role for the EHR to enable a precision-medicine approach for the treatment of recurrent kidney stone disease. A major strength of the EHR is the longitudinal data available across a large population with granular phenotyping data, such as stone composition and 24-hour urine data. The replication of *UMOD* from previous GWAS findings validates the EHR as a research data source and a clinical environment for utility testing. We additionally demonstrate that use of *UMOD* as a biomarker needs further evidence to support its clinical utility within the context of disease severity. Put differently, we show that while genetics may influence risk, that risk may not translate to measurable disease severity. Furthermore, we identify 19 novel risk loci for subgroups of by kidney stone composition, and the biological pathways associated with these variations need to be evaluated by functional genomics studies.

## Conclusions

In this EHR-based GWAS we replicate *UMOD* variants associating with kidney stone disease risk, but not disease severity. Stone composition and 24-hour urine studies comparing *UMOD* variants provide evidence supporting its role in urinary calcium crystallization. We identify novel genetic variants associated with specific subgroups of individuals with kidney stones as classified by stone composition, however the biologic pathways relating to risk need further elucidation. These findings suggest there may be a role for genetic testing linked to EHRs to facilitate precision-medicine approaches for the treatment of kidney stone disease.

## Data Availability

All data produced in the present work are contained in the manuscript.

## Acknowledgements

Supported by National Institutes of Health Grant R21DK127075 and CTSA award No. UL1 TR002243 from NCATS/NIH. Its contents are solely the responsibility of the authors and do not necessarily represent official views of the National Center for Advancing Translational Sciences or the National Institutes of Health.

